# COVID-19 County Level Severity Classification with Imbalanced Dataset: A NearMiss Under-sampling Approach

**DOI:** 10.1101/2021.05.21.21257603

**Authors:** Timothy Oladunni, Sourou Tossou, Yayehyrad Haile, Adonias Kidane

## Abstract

COVID-19 pandemic that broke out in the late 2019 has spread across the globe. The disease has infected millions of people. Thousands of lives have been lost. The momentum of the disease has been slowed by the introduction of vaccine; however, some countries are still recording high number of casualties. The focus of this work is to design, develop and evaluate a machine learning county level COVID-19 severity classifier. The proposed model will predict severity of the pandemic in a county into low, moderate, or high. Policy makers will find the work useful in the distribution of vaccines. Four learning algorithms (two ensembles and two non-ensembles) were trained and evaluated. Class imbalance was addressed using NearMiss under-sampling of the majority classes. The result of our experiment shows that the ensemble models outperformed the non-ensemble models by a considerable margin.

## I. INTRODUCTION

Since the outbreak of the coronavirus pandemic, the Centers for Disease Control and Prevention (CDC) has recorded close to 30 million cases. Thousands of lives have been lost to COVID-19 [1]. While the United States and other developed countries have been able to bend the curve on the fatality rate, emerging evidence suggests that the disease is just taking root in some countries. As of May 19, 2021, Mexico tops the fatality rate with 9.3%. At a distant second is Peru with 3.5%. Italy and Iran came third and fourth with 3% and 2.8% respectively [2]. The origin of this pandemic is an ongoing research; however, most scientists believe that it originated from a bat in Wuhan, China [3].

The question is: how do we categorize the severity of COVID-19 fatality in a county? We answered this question by building a machine learning classifier using the fatality rate dataset from the 3 006 counties in the US. Dataset was obtained from the John Hopkins University repository [2].

Machine learning algorithms have been shown to have the capability to learn pattern and discover knowledge from a dataset. It has been used in image recognition [4], fraud detection [5], voice recognition [6], malware detection [7], housing price prediction [8] etc. Since the outbreak of the coronavirus pandemic, several studies have been done using machine learning algorithms to understand the pandemic and provide strategies to reduce its spread.

Author [9] proposed a quantitative model to predict vulnerability to COVID-19 using genomes. Neural networks and Random Forests were used as learning algorithms. The result of the study confirmed previous work on phenotypic comorbidity patterns in susceptibility to COVID-19. In another study, Kexin studied nineteen risk factors associated with COVID-19 severity. The result suggested that severity relates to individual’s characteristics, disease factors, and biomarkers [10]. Hina et al., proposed a model to predict patient COVID-19 severity in Pakistan. Seven learning algorithms were trained and evaluated. The result of the experiment showed that Random Forest had the best performance with 60% accuracy.

While there are several studies on COVID-19 severity, there seems to be a gap in machine learning literature on the imbalanced classification of COVID-19 severity at the county level. Therefore, the focus of this study is the algorithmic imbalance classification of COVID-19 dataset of a county into low, moderate, or high. *We hypothesized that ensemble learning in conjunction with the NearMiss under-sampled majority class of an imbalance COVID-19 dataset has a superior capability of predicting the severity of COVID-19 at the county level*.

We tested our hypothesis by experimenting with ensemble and non-ensemble learning algorithms. Random Forest and Boosting Trees were trained and evaluated as our ensemble model, while Logistic Regression and K Nearest Neighbors as the non-ensemble models.

This paper is organized as follows: Section 2 describes the methodology for the study. Discussions, and conclusions are highlighted in sections 3 and 4 respectively. The limitation of the study was addressed in section 5. Finally, we acknowledged the source of our funding in section 6.

## II. METHOGOLOGY

The experimental flowchart of the study is shown in figure 1. The diagram shows the information flow of the proposed model. Fatality rate is a continues variable, therefore, categorization was done to convert it to discrete variables. Labeling was done into low, moderate, and high. Other variables in the dataset were processed as the predictive variables. Insignificant and redundant features were dropped during the cleaning phase. We also normalized the dataset. Four learning algorithm models were trained and evaluated. Performance comparison of the models was done using precision, recall, accuracy and F1-score.

**Figure 1.**
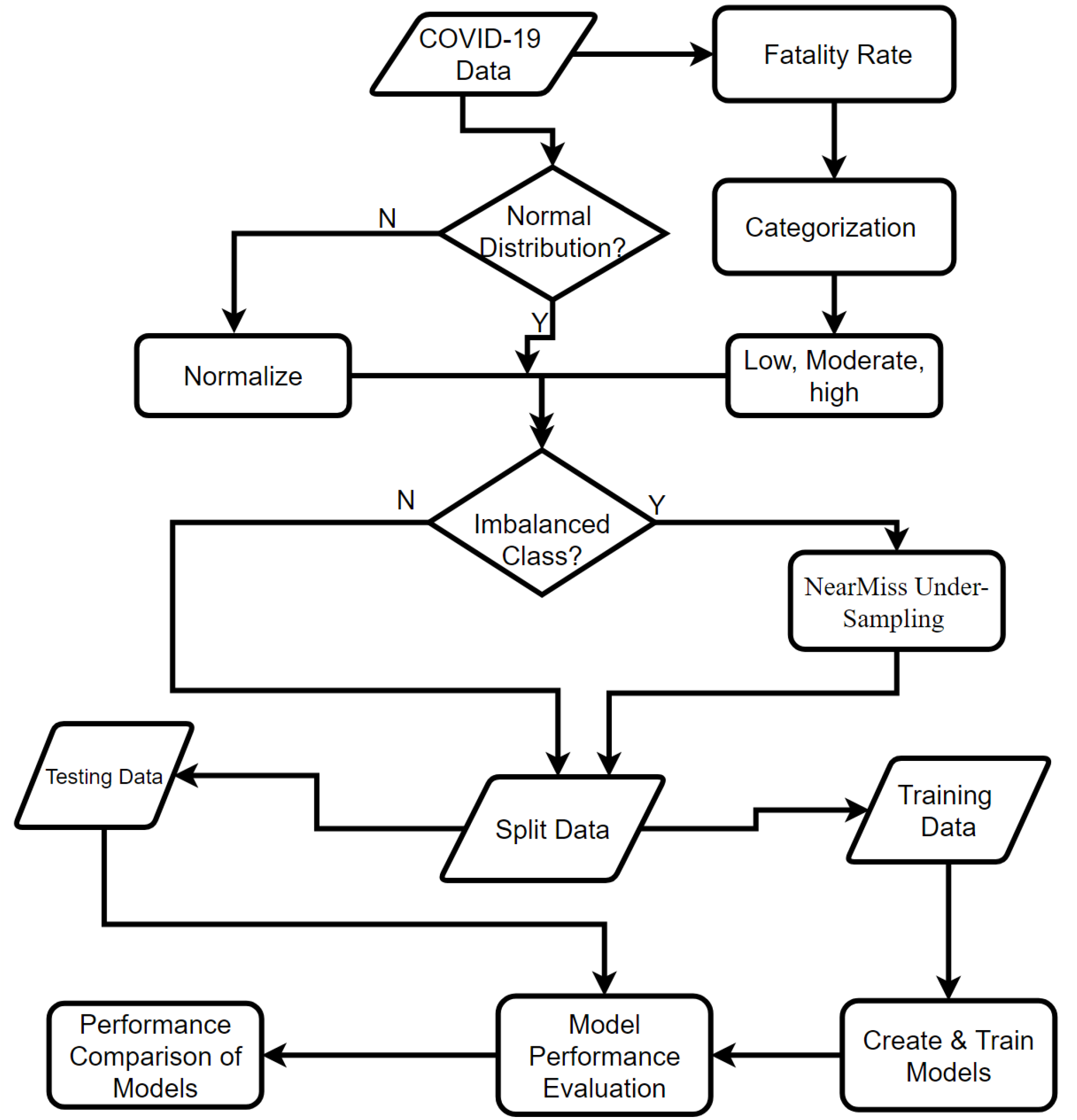
Experimental Flowchart.

### 1. Dataset

Dataset was obtained from the John Hopkins University COVID-19 repository [2]. Data consisted of the 3 006 counties of the United States. Dataset was cleansed at the processing stage.

### 2. Categorization

Severity of COVID-19 of a county was measured using the fatality rate as the response variable. Fatality rate attributes were split into 3 groups based on the following criterion: counties with fatality rates less than 1 were categorized as low (0 < x ≤ 1). Moderate class are the counties with fatality rate greater than 1 but less than or equal to 2 (1 < x ≤ 2). Finally, the high class are counties that have greater than 2 but less than equal to 4 fatalities (2 < x ≤ 4). Categorization or discretization is crucial for classification of continuous variables.

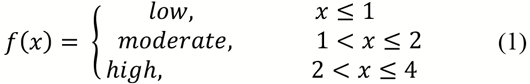

### 3. Imbalance Class

The above categorization resulted into skewed class distribution. This skewness of the class distribution is referred to in some literatures as class imbalance [11]. An imbalance dataset has one or more classes with low records (minority class) and one or more classes with many records (majority class). Class imbalance has been shown to have a considerable negative impact on the effectiveness of a learning algorithm [12].

### 4. Under-sampling of the majority class-The Near Miss Under-sampling (NMU) Approach

The question is, *how do we balance the dataset?* An imbalanced data can be balanced by oversampling of the minority class [13] or under-sampling of the majority class [14]. In oversampling approach, more data are created to increase the size of the minority class records to equal the majority class records. However, this approach has the risk of overfitting [15]. On the other hand, in under-sampling, the size of the majority class is reduced to balance the class distribution. This approach too has a tendency of underfitting the dataset.

Near Miss Under-sampling (NMU) appraoch was used in this study. NMU selection is based on distance of the majority class records to that of the minority class records [16]. It is a k nearest neighbor approach. The Euclidean distance can be used as the distance measure. NMU has three versions: version 1, version 2 and version 3 [17]. Version 1 is based on the smallest average distance between the majority class and three closest records of the minority class. Version 2 selects records from the majority class with farthest distance from three minority class. Lastly, in version 3, a given number of the majority class is selected for each closest example in the minority class. In this study, version 1 was used. The NearMiss function from the imblearn.under_sampling of the python library was imported. The result of our experiment showed the effectiveness of our strategy.

### 5. Experiment

We trained and evaluated 2 ensemble learning algorithms (Random Forest and Boosting trees). We also trained and evaluated 2 non-ensembles (Logistic Regression and K Nearest Neighbors). Dataset was split into 90% and 10% for training and testing, respectively.

#### 5.1 Performance Evaluation

To compare the results of our experiment, we used accuracy, precision, recall, and the F-1 score as comparison criteria.

##### 5.1.1 Accuracy

Accuracy is defined as the percentage of correct predictions for the test data. It can be calculated by dividing the number of correct predictions by the number of total predictions. The formula is as follow:

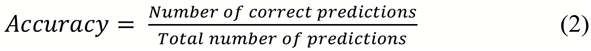

##### 5.1.2 Precision

Precision is a metric that quantifies the number of correct positive predictions made. It is calculated using the following formula:

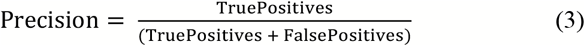

##### 5.1.3 Recall

Recall is a metric that quantifies the number of correct positive predictions made from all positive predictions that could have been made. Its operation is as followed:

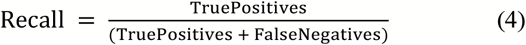

##### 5.1.4 F-Measure

F-Measure provides a way to combine both precision and recall into a single measure that captures both properties. Its formula is

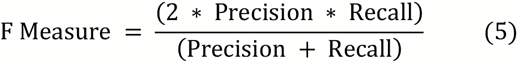

#### 5.2 Training and Performance Evaluation Learning Algorithms

We trained and evaluated the performances of 4 learning algorithm.

##### 5.2.1 K-Nearest Neighboring (KNN)

In a dataset with response variable *y* and **X** feature vectors, a KNN learning algorithm identifies K points in the training dataset that are closest to a new testing datapoint *x*_*0*_.

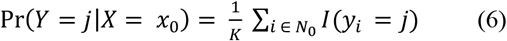

Where *j* is estimated response and *y*_*i*_ as the target (label). *N*_*0*_ are the K points. In our experiment, 5 was selected as the value of K. In addition, we used the MixedMeasures for the measure types. The Euclidean distance was used as the distance metric. [18]

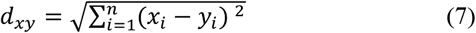

Where **d** represents the distance, **x** and **y** are 2 data points.

Performance of the KNN learning algorithm is shown in table 1. In all evaluation criterion, the result suggest that moderate class has the lowest prediction. Overall model accuracy score was approximately 0.61.

**Table 1.** KNN performance

| KNN TABLE |  |  |  |  |
| --- | --- | --- | --- | --- |
|  | classification_report |  |  |  |
|  | precision | recall | f1-score | support |
| High | 0.61 | 0.77 | 0.68 | 235 |
| low | 0.68 | 0.71 | 0.69 | 222 |
| moderate | 0.47 | 0.27 | 0.35 | 172 |
| accuracy |  |  | 0.61 | 629 |
| macro avg | 0.59 | 0.59 | 0.57 | 629 |
| weighted avg | 0.60 | 0.61 | 0.59 | 629 |
| confusion_matrix |  |  |  |  |
| [[181 23 31] |  |  |  |  |
| [ 42 158 22] |  |  |  |  |
| [ 72 53 47]] |  |  |  |  |
| accuracy_score 0.6136724960254372 |  |  |  |  |

##### 5.2.2 Logistic Regression

Logistic regression is a supervised learning algorithm for predicting the likelihood of a target variable. In a two-class problem, the target or dependent variable is dichotomous, which implies there would be just two potential classes [19]. The logistic function produces output between 0 and 1.

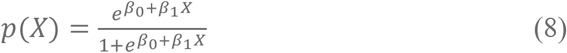

It can be shown that,

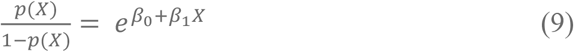

Taking logarithms,

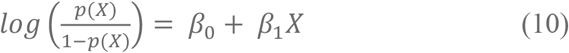

where β_0_ is the bias or intercept term and β_1_ is the coefficient for the single input value (x). L2 regularization was used as the overfitting control. Tolerance for stoppage criteria was 1e-4. Optimization was based on lbfgs (Limited-memory Broyden–Fletcher–Goldfarb– Shanno). Table 2 shows the result of the Logistic Regression.

**Table 2.** Logistic Regression performance.

| LOGISTIC REGRESSION TABLE |  |  |  |  |
| --- | --- | --- | --- | --- |
|  | classification_report |  |  |  |
|  | precision | recall | f1-score | support |
| High | 0.00 | 0.00 | 0.00 | 235 |
| low | 0.35 | 1.00 | 0.52 | 222 |
| moderate | 0.00 | 0.00 | 0.00 | 172 |
| accuracy |  |  | 0.35 | 629 |
| macro avg | 0.12 | 0.33 | 0.17 | 629 |
| weighted avg | 0.12 | 0.35 | 0.18 | 629 |
| confusion_matrix |  |  |  |  |
| [[ 0 235 0] |  |  |  |  |
| [ 0 222 0] |  |  |  |  |
| [ 0 172 0]] |  |  |  |  |
| accuracy_score 0.35294117647058826 |  |  |  |  |

As shown in table 2, with a model performance accuracy of 35.3%, the performance of Logistic Regression is worse than that of KNN.

##### 5.2.3 Random Forest

Random forest is a supervised learning algorithm that is utilized for classifications as well as regression. As a forest, Random Forest comprises of several decision trees. Intuitively, more trees suggest a stronger forest. Therefore, aggregating decision trees in ensemble learning, produces a better performance. Essentially, a Random Forest model computation is based on decision trees using bootstrapped training data samples.

The ensemble methodology of the Random Forest makes its prediction superior to that of a single decision tree; Random Forest decreases over-fitting by averaging the outcome of trees [20]. Using a randomized subset of the predictive variables ensures that prediction is not dominated by the most influential predictive variables; all predictive variables are given a chance. This arrangement leads to a decorrelating of the trees, thereby providing a substantial reduction in variance over bagging or a single decision tree. Table 3 shows the performance of the Random Forest model.

**Table 3.** Random Forest performance.

| RANDOM FOREST REGRESSION TABLE |  |  |  |  |
| --- | --- | --- | --- | --- |
|  | classification_report |  |  |  |
|  | precision | recall | f1-score | support |
| High | 0.52 | 0.91 | 0.66 | 235 |
| low | 0.78 | 0.77 | 0.78 | 222 |
| moderate | 0.00 | 0.00 | 0.00 | 172 |
| accuracy |  |  | 0.61 | 629 |
| macro avg | 0.43 | 0.56 | 0.48 | 629 |
| weighted avg | 0.47 | 0.61 | 0.52 | 629 |
| confusion_matrix |  |  |  |  |
| [[213 22 0] |  |  |  |  |
| [ 50 172 0] |  |  |  |  |
| [145 27 0]] |  |  |  |  |
| accuracy_score 0.6120826709062003 |  |  |  |  |

Table 3 shows that the Random Forest model outperformed KNN and Logistic Regression models.

##### 5.2.4 Boosting Tree

As shown in Table 3, compared to Logistic Regression and KNN, Random Forest showed some improvement, however, at a model accuracy of 61.2%. Therefore, we continued our investigation by exploring the Boosting Tree Algorithm. Just like Random Forest, Boosting is an ensemble modeling technique for creating a strong classifier from several decision trees. This is done by cascading weak models in series. First and foremost, a decision tree is built from the training data. The next model was fitted into the residual of the present model. This sequential learning continued, until either the total training data is predicted accurately, or the most extreme number of models are added [21]. The Boosting Algorithm is a slow learner. It has been shown that slow learning algorithms perform better than the fast ones [22].

Table 4 shows the performance outcome of the Boosting Tree.

**Table 4.** Boosting Tree Performance

| BOOSTING TABLE |  |  |  |  |
| --- | --- | --- | --- | --- |
|  | classification_report |  |  |  |
|  | precision | recall | f1-score | support |
| High | 0.96 | 0.97 | 0.97 | 235 |
| low | 0.96 | 0.98 | 0.97 | 222 |
| moderate | 0.94 | 0.90 | 0.92 | 172 |
| accuracy |  |  | 0.95 | 629 |
| macro avg | 0.95 | 0.95 | 0.95 | 629 |
| weighted avg | 0.95 | 0.95 | 0.95 | 629 |
| confusion_matrix |  |  |  |  |
| [[229 1 5] |  |  |  |  |
| [ 0 217 5] |  |  |  |  |
| [ 9 9 154]] |  |  |  |  |
| accuracy_score 0.9538950715421304 |  |  |  |  |

As shown in Table 4, the Boosting Tree Model showed a significant improvement over the three previous models.

## III. DISCUSSION

Accuracy of the models were compared. For each model, we also took the average performance of the precision, recall and F1 score. Table 5 shows the comparison table.

**Table 5.**
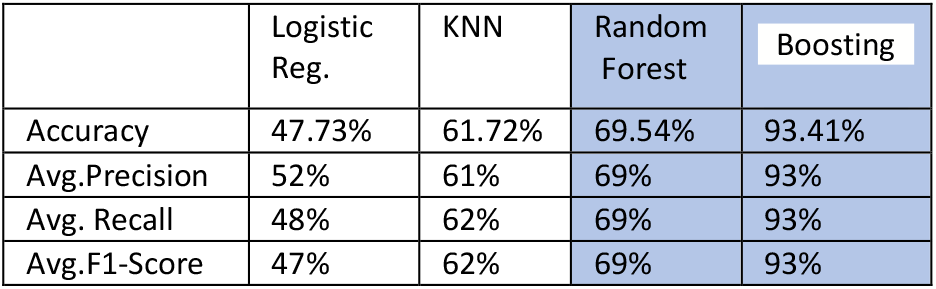
Model Performance Comparison

|  | Logistic Reg. | KNN | Random Forest | Boosting |
| --- | --- | --- | --- | --- |
| Accuracy | 47.73% | 61.72% | 69.54% | 93.41% |
| Avg.Precision | 52% | 61% | 69% | 93% |
| Avg. Recall | 48% | 62% | 69% | 93% |
| Avg.F1-Score | 47% | 62% | 69% | 93% |

As shown in the experimental result, the non-ensemble algorithms of Logistic Regression and KNN have the worst performance. The table also that the ensemble algorithms of Random Forest and Boosting Tree models outperformed other models. As discussed, these two models were built with large number of decision trees on bootstrapped training data. Boosting and Random Forest models have the best performances with 93.41% and 69.54% of accuracy respectively. Performances based on precision, recall and F1, the Boosting Model showed an averaged value of 93%, 93%, and 93% respectively.

The superior performance of the Boosting Model is not surprising because, a boosting tree is a large combination of decision trees grown sequentially. Random Forest and Boosting Tree are built on the ensemble of decision trees. However, the arrangement of fitting small trees with a few terminal nodes into the residual of the previous tress in a Boosting Tree sequentially improves the performance of the model.

## IV. CONCLUSION

In this study we have designed, developed, and evaluated a COVID-19 severity classifier using imbalance class dataset. The proposed model has the capability of predicting the severity level of COVID-19 in a county. Dataset was obtained from the JHU COVID-19 repository. COVID-19 Severity level was based on fatality rates in all the 3 006 counties of the US. For classification purpose, COVID-19 severity was categorized into low, moderate, and high.

Imbalance class was addressed using the Near Miss Under-sampling (NMU) approach. Ensemble and non-ensemble learning algorithms were trained and evaluated. Ensemble models include Random Forest and Boosting Trees. KNN and Logistic Regression were used as the non-ensemble models.

The result of our experiment suggests that the ensemble models in conjunction with NMU are the most effective in building a COVID-19 severity classifier at the county level using imbalanced dataset. Thus, we do not have sufficient evidence against our hypothesis. Therefore, we contend that *ensemble learning in conjunction with the NMU under-sampled majority class of an imbalanced COVID-19 dataset has a superior capability of classifying the severity of COVID-19 at the county level*.

## V. LIMITATION OF STUDY

Dataset for the study contained the 3 006 counties of the United States. As shown in the experiment, fatality rate was discretized into low, moderate, and high. Class obtained was imbalanced. Since the impact of COVID-19 is different in various countries, severity classes may be different. Therefore, this study may have a different outcome.

## Data Availability

John Hopkins University COVID-19 Repository

## VI. ACKNOWLEDGEMENT

This work is funded by the National Science Foundation grant number 2032345.

